# Efficacy of an Automated Point-of-Care UVC Disinfection System for Reusable Ophthalmic Devices

**DOI:** 10.1101/2025.09.15.25335810

**Authors:** Vikram A. Shankar, Aakriti G. Shukla, Phillip Ianchulev, Alan L. Robin, David F. Chang

## Abstract

**Purpose:** Disinfection of multi-use ophthalmic equipment prevents cross-transmission of microbes between patients. This study evaluates the germicidal efficacy of the Saniteyes^TM^ ultraviolet-C (UVC) high-level disinfection system for multi-use ophthalmic equipment such as eyedropper bottles, tonometer tips, and diagnostic and laser lenses.

**Methods:** We assessed disinfection efficacy using both carrier and simulated use tests against multiple bacterial, viral, and fungal microorganisms. Organisms were selected based on commonly accepted indicators for disinfection efficacy and/or ophthalmic relevance. Stainless steel carriers, eyedropper nozzles, applanation prisms, goniolenses, and laser lenses were inoculated before being placed within a disinfection bay of the Saniteyes^TM^ system. Following disinfection, the log reduction in viable organisms was measured. Tests were performed according to American Society of Testing and Materials (ASTM) International Standards for high-level disinfection (HLD). All tests were performed with negative controls, soiled controls, and three replicates for efficacy studies.

**Results:** Carrier and simulated use tests showed reduction by more than 6-log for all bacteria and fungi, and by more than 4-log for viruses, in accordance with efficacy cutoffs for HLD.

**Conclusions:** The Saniteyes^TM^ automated disinfection system offers fast and chemical-free HLD of multi-use ophthalmic devices to meet regulatory standards of the CDC, FDA, and Joint Commission.

## Introduction

Healthcare-associated infections affect approximately 2 million patients annually in the United States and result in nearly 100,000 deaths per year.^1^ In response, regulatory agencies such as the Food and Drug Administration (FDA) and The Joint Commission (TJC) have increased scrutiny of reusable medical devices for their potential role in cross-transmission. These agencies establish disinfection standards using the century-old Spaulding classification system, which categorizes medical devices based on their invasiveness and potential for patient harm.^2,3^ Under this framework, many ophthalmic devices are classified as *semi-critical* due to their contact with the ocular surface and must undergo high-level disinfection (HLD) to eliminate all living organisms before reuse.^4^ Originally formulated for invasive equipment like endoscopes and laryngoscopes, these regulations have created implementation challenges in ophthalmology, where reusable devices are employed in over 100 million eye examinations each year in fast-paced clinical settings.^4,5^

In 2019, The Joint Commission reinforced HLD mandates for tonometry tips, goniolenses, and laser lenses following outbreaks of epidemic keratoconjunctivitis (EKC) and herpes simplex virus (HSV) in eye clinics.^3,4,6^ Chemical HLD with bleach or enzymatic agents remains the gold standard but is variably followed in outpatient settings due to time constraints, labor, and uncertain value.^7,8^ Larger institutions including academic centers circumvent HLD requirements by switching to single-use disposable devices, increasing environmental waste and expense.^9^ Multi-use eyedropper bottles are classified as drug-device combination products by the FDA but remain outside the purview of HLD mandates. Bottles are approved for reuse until expiration despite paradoxically lacking any methods of disinfection between patients.^10,11^ While microbial contamination of the nozzle tip and cross-transplantation may be benign for patients with a healthy ocular surface, it can pose greater risk for patients undergoing intravitreal injections or surgical procedures.^12,13^

A more practical disinfection method is needed that balances the imperatives of patient safety and sustainability with the realities of high-volume clinical ophthalmology. To address this unmet need, an automated point-of-care disinfection device (Saniteyes^TM^, Dropmate, Inc., Sunnyvale, CA) was developed to provide rapid, consistent, and effective reprocessing of reusable ophthalmic instruments (**Figure 1**). This compact disinfection system employs germicidal irradiation with primary reliance on ultraviolet-C (UVC) light to inactivate bacterial, viral, and fungal pathogens on eyedropper bottles and other reusable semi-critical diagnostic equipment. UV germicidal irradiation (UVGI) has been validated across diverse healthcare environments as a fast, reliable, and chemical-free alternative to conventional disinfection.^14–18^ The Saniteyes^TM^ device is the first FDA-cleared technology that uses light-based disinfection to reprocess semi-critical ophthalmic instruments, and the first device to our knowledge capable of disinfecting in-use eyedropper bottles.

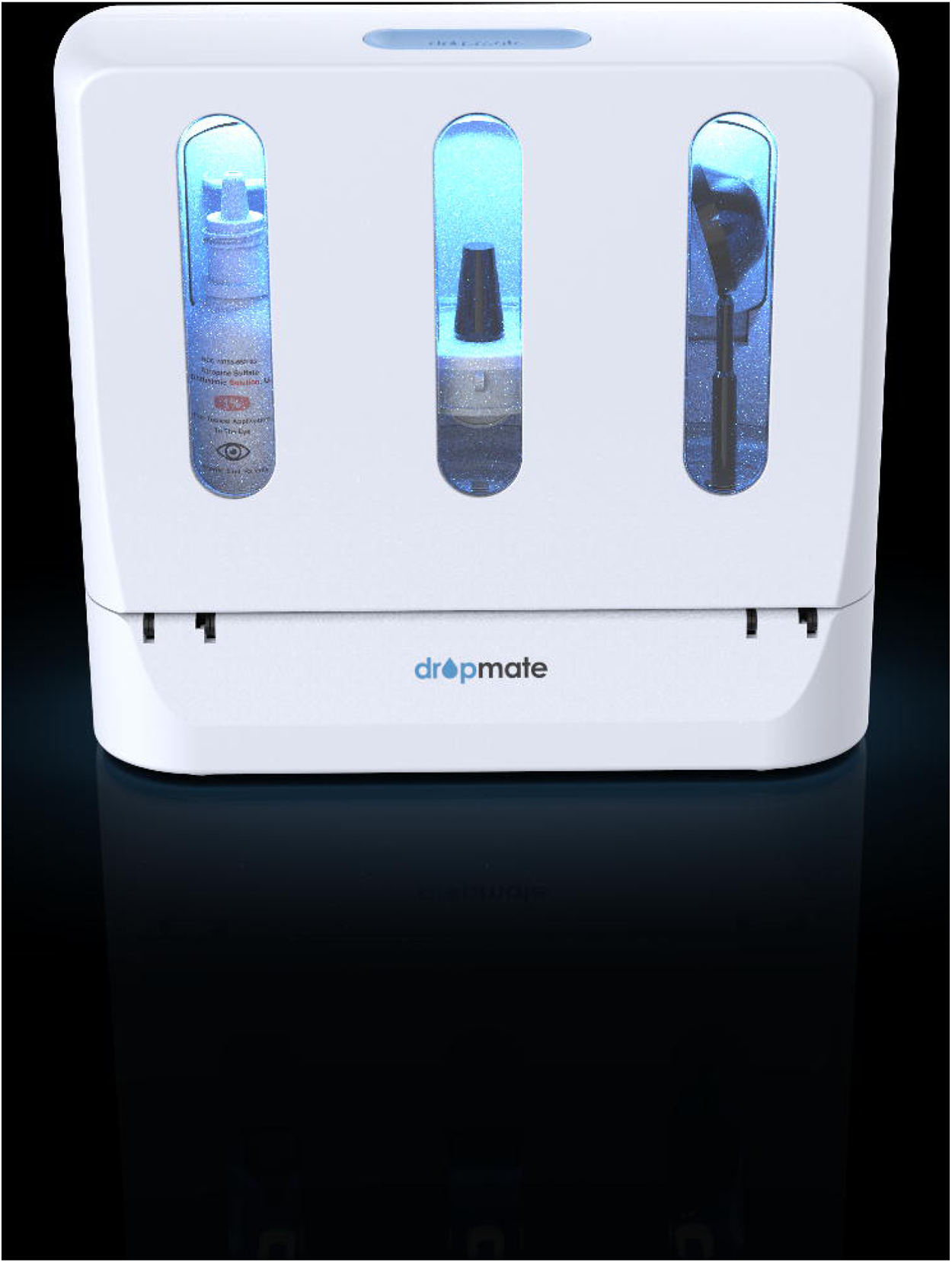

## Materials and Methods

The Saniteyes^TM^ disinfection system evaluated in this study delivers UVC light with wavelengths between 254–280 nm to induce pyrimidine dimer formation in microbial DNA and RNA, disrupting replication and inactivating pathogens in a dose-dependent fashion.^14,15^ Photochromic indicator strips (Intellego Technologies, Solna, Sweden) were used to verify sufficient levels of germicidal irradiation to achieve HLD endpoints of 6-log reduction of bacteria and fungi and 4-log reduction of viruses. These strips undergo a predictable color change in response to UVC light, providing a visual, real-time assessment of radiant fluence (mJ/cm^2^) delivered to a target surface. They offer a simple, reproducible, and effective method for validating disinfection protocols for health care applications.

Following dose validation, microbiological testing was carried out using both inoculated carriers and simulated use tests with commonly used ophthalmic equipment. A range of organisms was chosen based on their use as established indicators of disinfection efficacy, with particular attention paid to pathogens of ophthalmic significance. All carrier and simulated use tests were run using standard 5- and 10-minute processing cycles. Efficacy testing was independently conducted by Prime Analytical Labs, LLC (Concord, CA, USA). Bactericidal tests were conducted according to AOAC Official Methods 991.48 and 991.49 against Staphylococcus aures and Pseudomonas aeruginosa. Mycobactericidal tests were carried out according to AOAC Official Method 965.12 against Mycobacterium terrae. Fungicidal tests were carried out according to AOAC Official Method 955.17 against Candida albicans.

### Carrier tests

Six stainless steel 1” round carriers were inoculated with 0.1 mL of each challenge organism at a concentration of >10^6^ CFU/0.1mL and allowed to visibly dry. Each carrier was placed into the Saniteyes^TM^ disinfection device following manufacturer instructions and exposed for 5 or 10 minutes. For bacterial and fungal tests, carriers were removed from the device and placed into a sterile plastic container with 10mL of phosphate buffered saline (PBS). PBS. An un-exposed, control coupon was prepared in the same fashion. The coupons were massaged with the PBS to recover the spiked culture, and from each plastic container a 10-fold serial dilution was prepared and labeled. 100uL from each solution was aseptically pipetted onto corresponding labeled plates and spread. Each step was repeated for each challenge organism to complete all designated exposure timepoints. An inoculum verification (IV) sample was also prepared from each challenge organism suspension using the same method. All tests were performed in triplicate. For viral tests, coupons were removed from the device following disinfection and placed into sterile plastic containers containing 1mL of viral recovery medium. Eluates were collected and used to perform a TCID_50_ infectivity assay using the Reed-Muench method. All tests were performed in triplicate.

### Simulated use tests

Simulated use tests were carried out on representative ophthalmic devices—eyedropper bottles, Goldmann 3-mirror goniolenses, multi-dose drug vials, and applanation prisms (**Figure 3**). Each device was disinfected before inoculation by soaking in 10% dilute bleach for 10 minutes, followed by rinsing and air drying. Eyedropper nozzles and the contact surfaces of each goniolens and applanation prism were spiked with 0.1 mL of the challenge organism at a concentration of >10^6^ CFU/0.1mL and allowed to visibly dry. To match HLD standards in the United States and European Union for tonometer tips and goniolenses, a mechanical cleaning step was performed before irradiation using a common 70% isopropyl alcohol wipe. Each device then was placed into the Saniteyes^TM^ disinfection device following manufacturer instructions and exposed for specified time durations of both 5 and 10 minutes. The exposed devices were removed from the disinfection system and placed into a sterile plastic container with 10mL of PBS. Un-exposed, control devices were prepared in the same fashion and placed into a sterile plastic container with 10mL of PBS. The devices were massaged with the PBS to recover the spiked culture, and from each plastic container a 10-fold serial dilution was prepared and labeled. From each dilution, 100uL was aseptically pipetted onto corresponding labeled plates and spread with a hockey stick. Each step was repeated for each challenge organism to complete all designated exposure timepoints. An inoculum verification (IV) sample was also prepared from each challenge organism suspension using the same method. All tests were performed in triplicate.

The plates were prepared and incubated according to the conditions in Table 1. At the end of the incubation period, the CFUs were counted for each dilution and recorded accordingly. For each dilution of the positive control and inoculum verification, the plate count was performed and recorded. The log reduction was calculated using the formula: Log Reduction = log10 (initial CFU / final CFU).

**Table 1.**
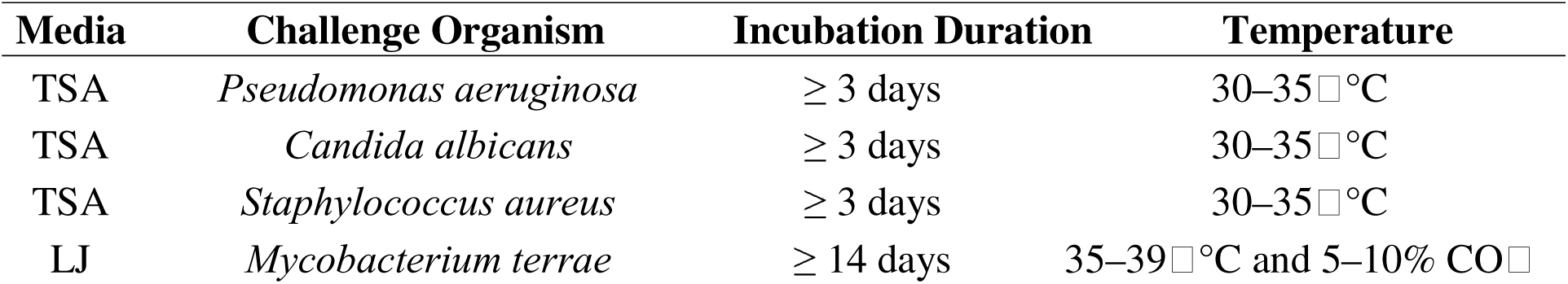
Media/Organism Incubation Conditions.

## Results

### UVC Exposure tests

Exposure tests were performed according to manufacturer instructions and are presented in **Figure 2**. Photochromic indicators at multiple positions on each eyedropper nozzle confirmed consistent and uniform saturation of all potential contact surfaces across all disinfection bays and positions (**Table 2**). Cumulative dose reached or exceeded 80 mJ/cm^2^ across every tested position in each bay and was verified using the dosimeter reference card provided by the manufacturer shown in **Figure 3**.

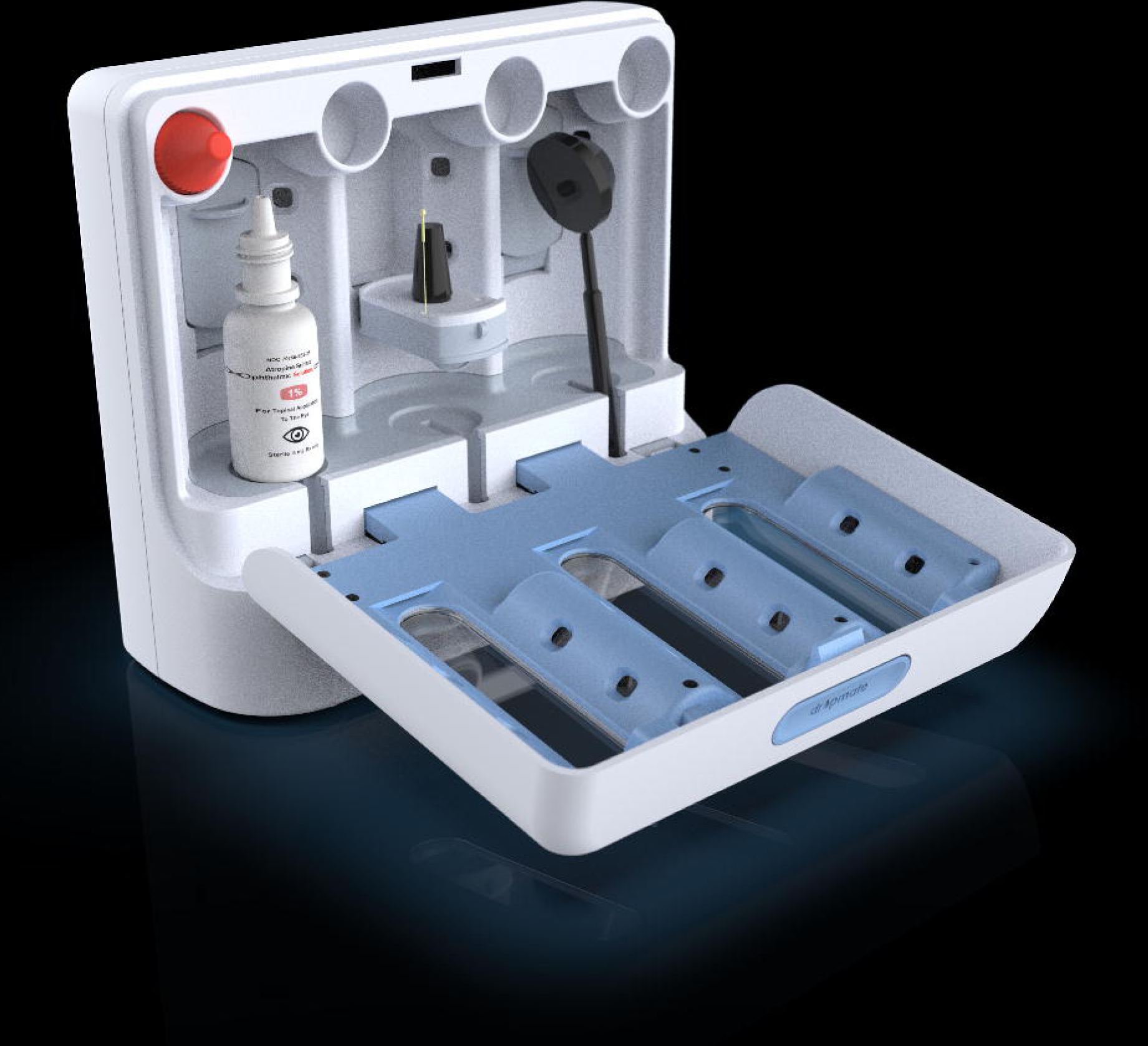

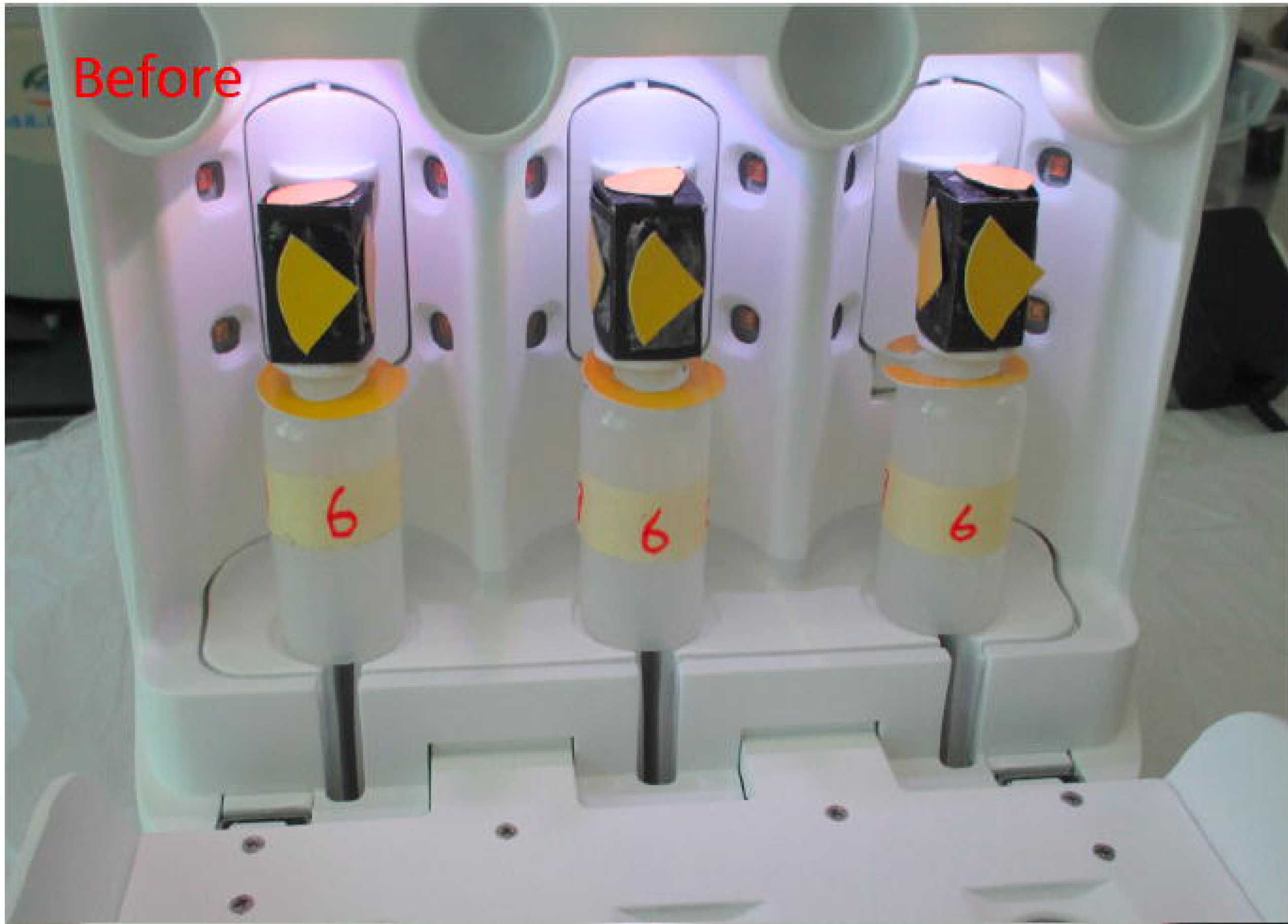

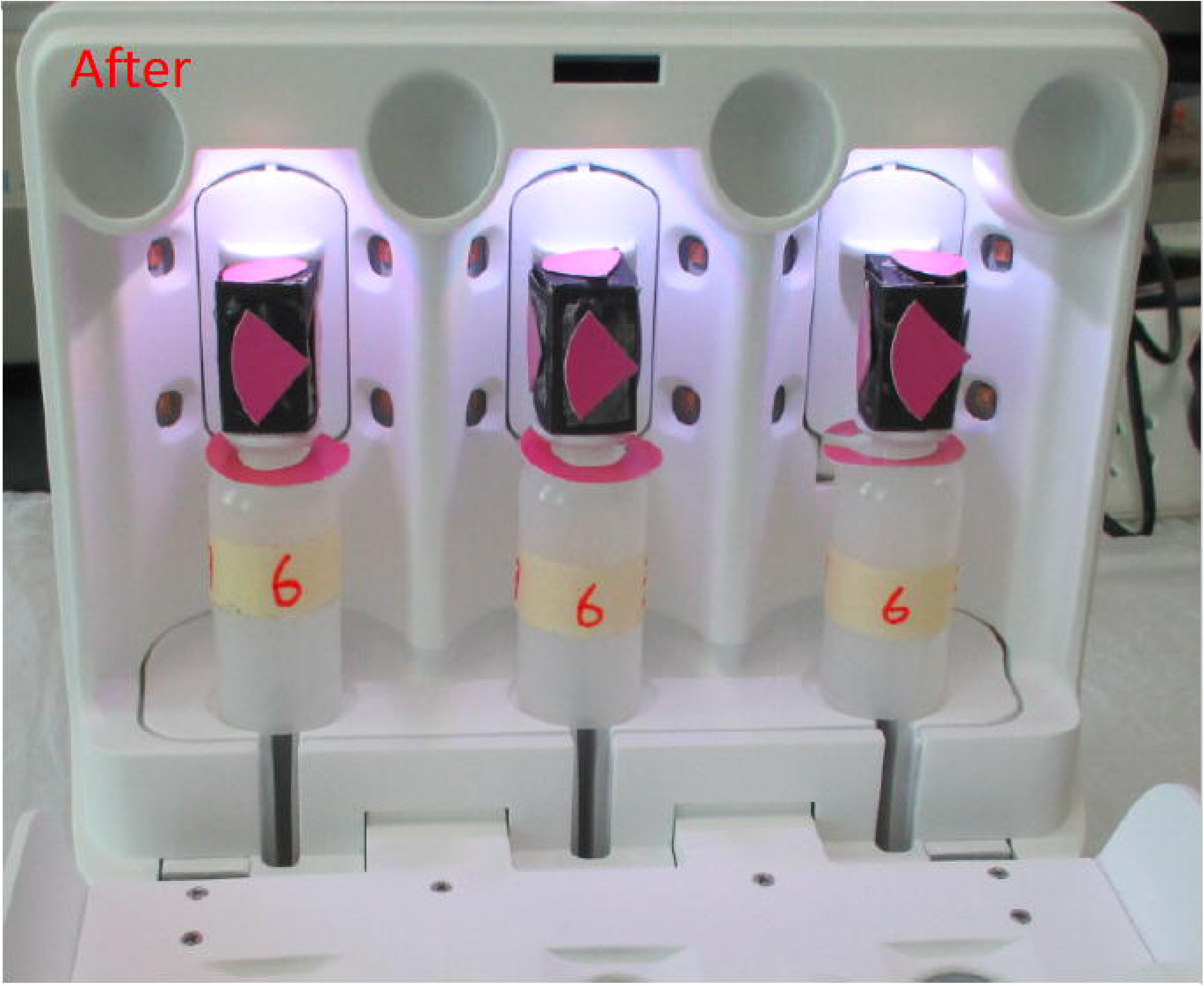

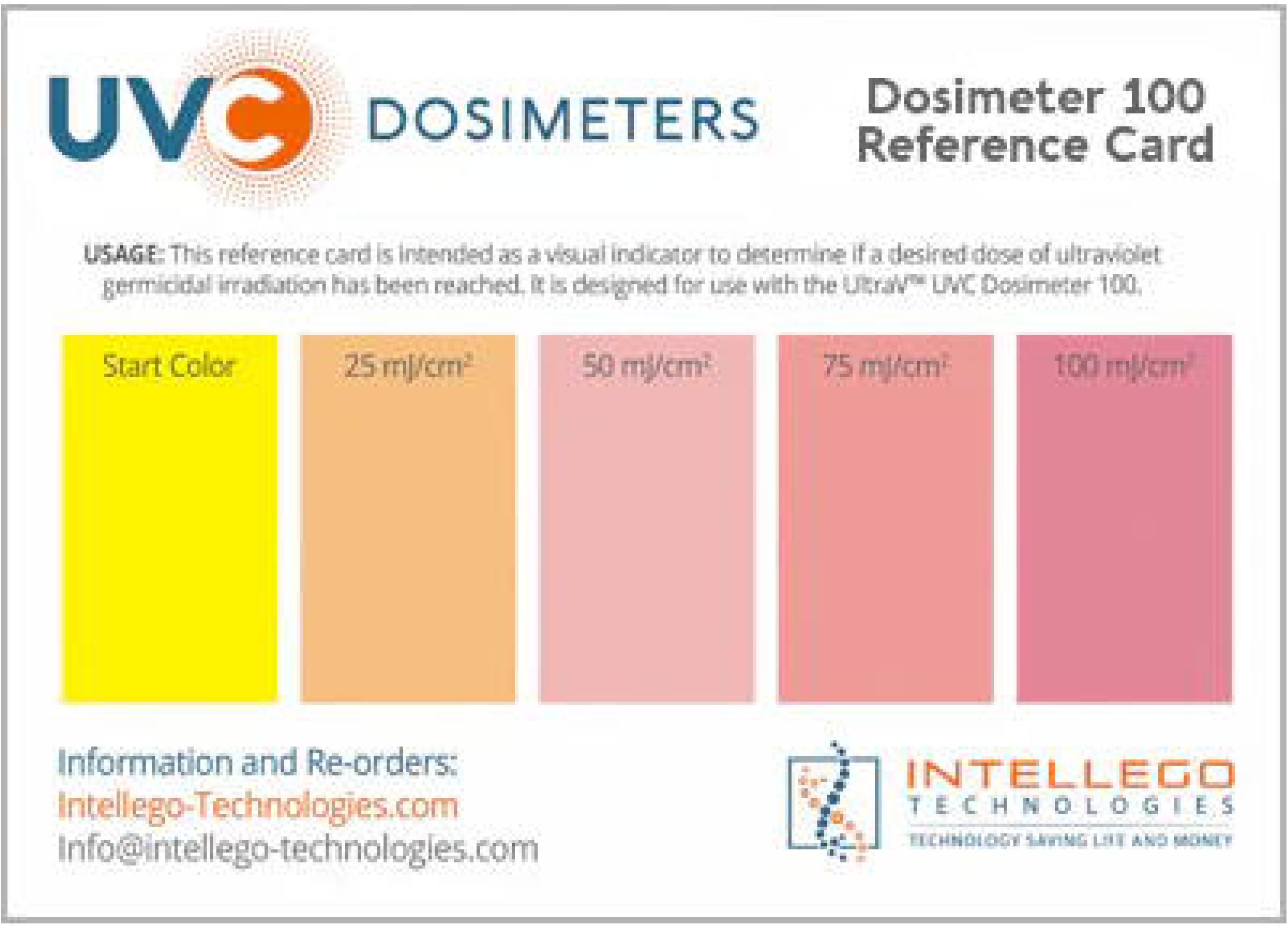

**Table 2.**
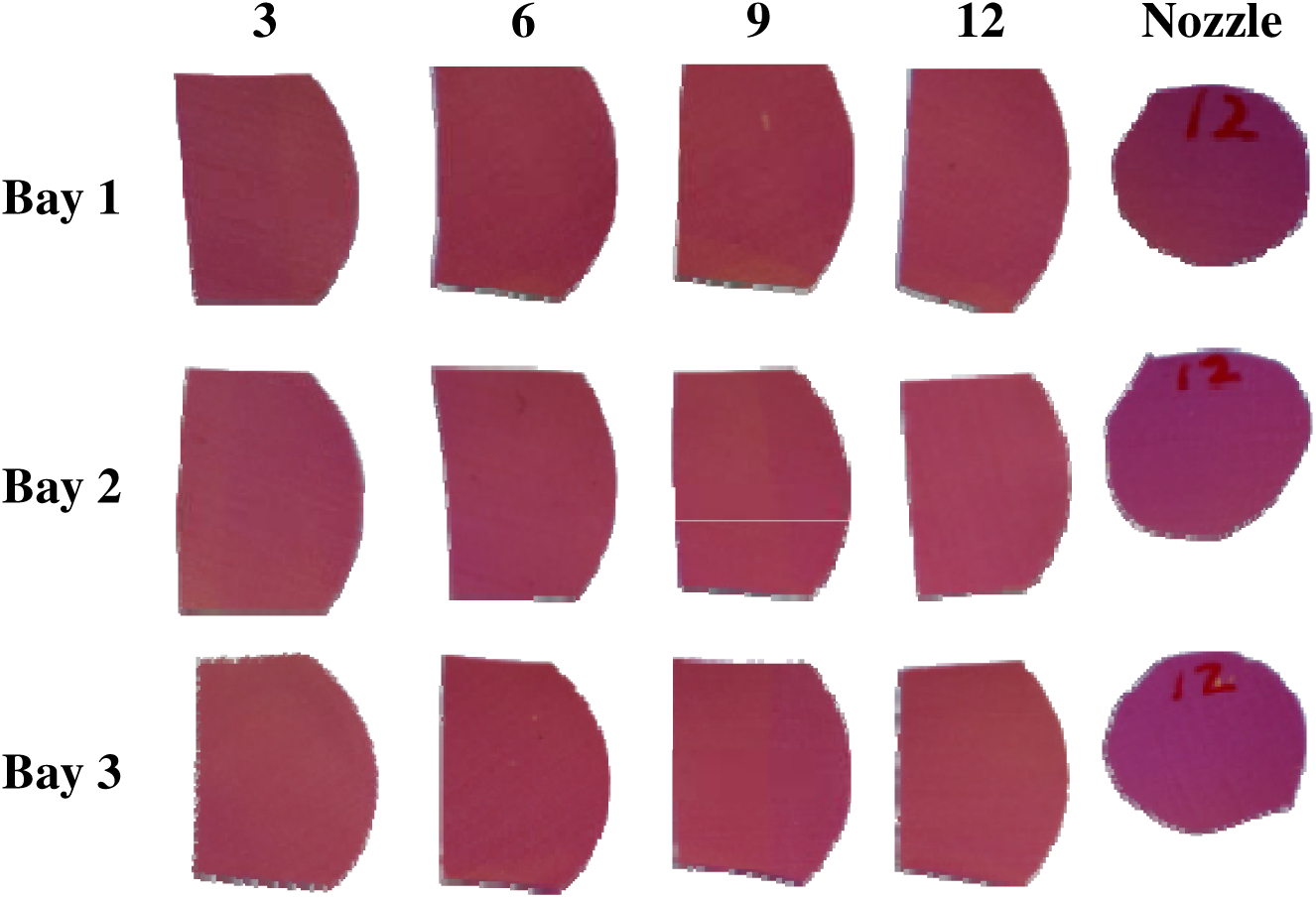
Cumulative UV-C exposure by position with reference scale. UV-C colorimetric indictors were placed in three bays (rows) at four positions (columns labeled 3,6,9 and 12 o’clock) and on the nozzle tip to assess cumulative exposure. Color change reflects total UV-C dose delivered, with consistent and uniform saturation observed across all disinfection bays and positions without cold spots.

### Carrier tests

Carrier test results are presented in **Table 3**. All controls were within normal ranges and all samples passed the efficacy cut-offs for HLD. The mean log reduction in organism load was calculated for each of the carrier test cycles. All controls were within normal ranges, and all samples tested passed efficacy cut-offs of 6-log reduction of bacteria and fungi, and 4-log reduction of viruses. These data indicate the device satisfies the criteria for HLD under both EN and ASTM standards.

**Table 3.**
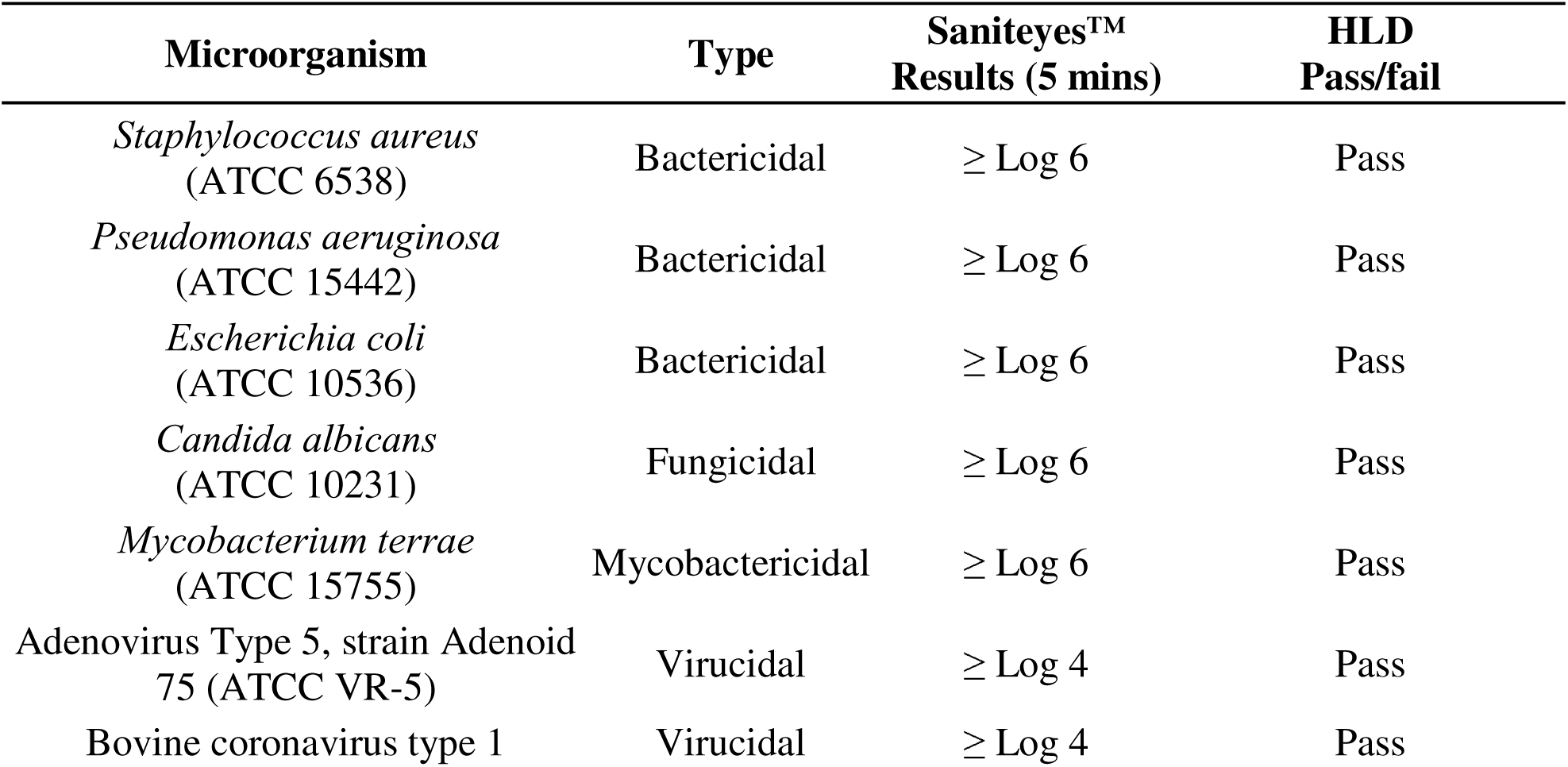
Carrier Tests.

### Simulated use tests

The results of tests simulating worst-case conditions are shown in **Table 4**. The mean log reduction in viable organism count and the SEM were calculated for each simulated use test. The mycobacterial load was reduced by more than 6-log in all cases. These results indicate that the device is capable of HLD under simulated use conditions and meets efficacy cutoffs established by regulatory bodies like the CDC, FDA, and TJC.

**Table 4.**
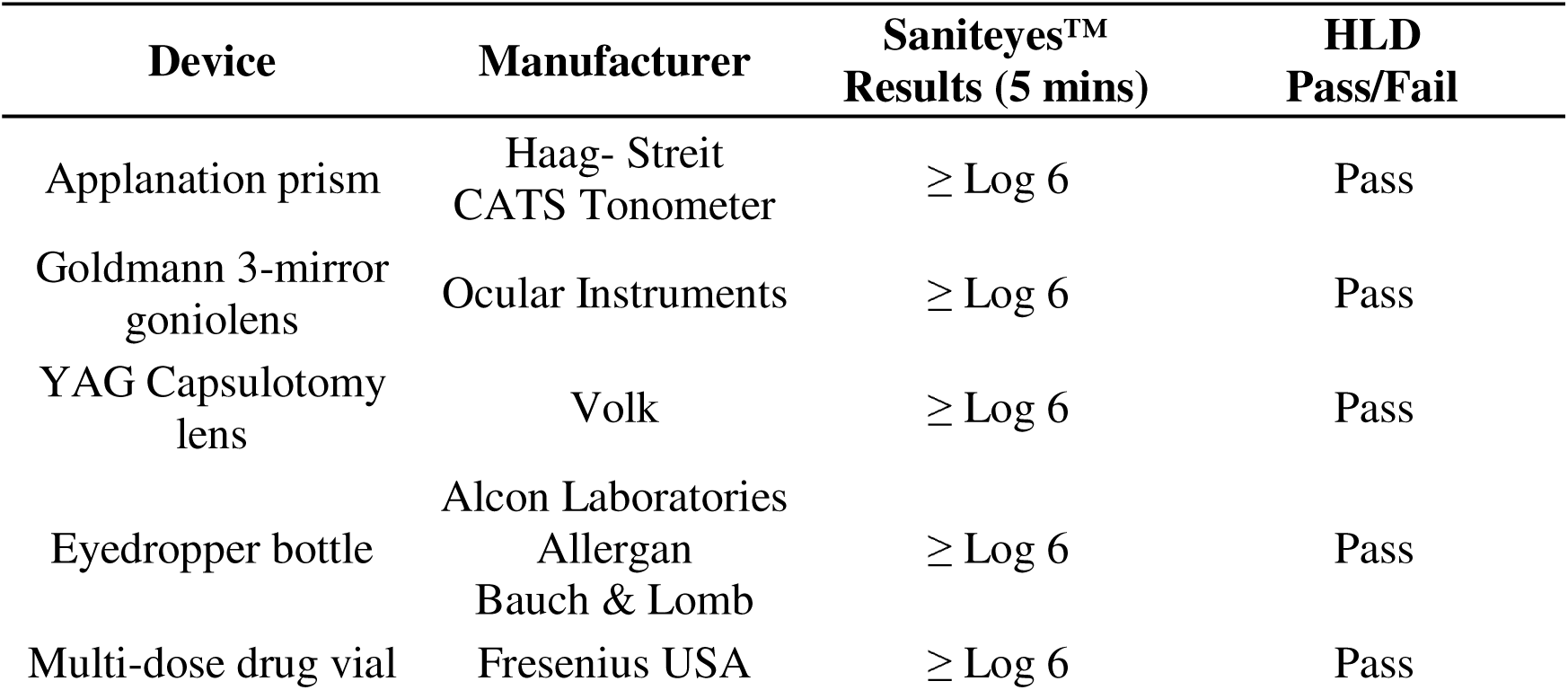
Mycobacterial Simulated Use Tests.

| **Device** | **Manufacturer** | **Saniteyes™**<br>**Results (5 mins)** | **HLD**<br>**Pass/Fail** |
| --- | --- | --- | --- |
| Applanation prism | Haag- Streit<br>CATS Tonometer | ≥ Log 6 | Pass |
| Goldmann 3-mirror<br>goniolens | Ocular Instruments | ≥ Log 6 | Pass |
| YAG Capsulotomy<br>lens | Volk | ≥ Log 6 | Pass |
| Eyedropper bottle | Alcon Laboratories<br>Allergan<br>Bauch & Lomb | ≥ Log 6 | Pass |
| Multi-dose drug vial | Fresenius USA | ≥ Log 6 | Pass |

## Discussion

The FDA-cleared Saniteyes^TM^ automated disinfection system consistently achieved HLD standards in both carrier and simulated use tests with eyedropper bottles, multi-dose drug vials, diagnostic and laser lenses, and tonometer tips. Photochromic indicator strips confirmed uniform delivery of UVGI with the elimination of shadowing or cold spots that can harbor microorganisms after reprocessing. The testing methodologies and microbial bioburdens in this study were deliberately more challenging than would be encountered in real-world clinical scenarios, validating the efficacy of the Saniteyes^TM^ system for HLD of reusable ophthalmic devices in “worst case” scenarios.

Although FDA-approved UVGI reprocessing systems exist across health care for devices like endoscopes and ultrasound probes, ophthalmology continues to rely on legacy chemical disinfection methods.^19,20^ Agents like bleach and hydrogen peroxide require significant manual labor and up to ten minutes for reprocessing, which can be impractical in fast-paced clinical settings. Chemicals carry a risk of iatrogenic corneal toxicity if not rinsed off before reuse and can shorten the service life of instruments by degrading plastic, dissolving adhesives, and impairing lens optics.^3,4^ Chemical HLD protocols are also limited by a lack of standardization, as approved agents and immersion times vary among regulators, manufacturers, and professional societies. ^3^ In practice, less than 10% of surveyed practices use chemical HLD methods due to time constraints, labor, or inconvenience, leaving a gap in the standard of care.^8^

Large, high-volume institutions like the VA Hospital System and academic centers prefer to circumvent HLD requirements by transitioning to single-use disposable plastics.^4,8^ Single-use instruments have gained favor following the COVID-19 pandemic despite recent studies questioning their financial and environmental costs.^21^ A large institution may spend more than $1 million over a decade switching to single-use goniolenses and applanation tips, with cost-effectiveness ratios greater than $61,000 per case of EKC averted.^9,21,22^ The environmental impact is also significant, as a single outpatient department can generate more than 110 kg of plastic waste each year using single-use tonometer prisms and goniolenses.^23^ Despite the perceived safety of single-use, half of all technicians and staff acknowledge accidentally touching the contact surface of disposable tonometer prisms during, compromising sterility.^24^ As health care reckons with its growing cost and environment footprint, sustainable disinfection and reuse of equipment should be prioritized.^10,11^

While tonometer tips and lenses must undergo HLD under current regulations, multi-use eyedropper bottles operate under different guidelines that permit reuse despite lacking any disinfection methods.^11^ Preservatives inhibit microbial growth in residual solution but contamination of the nozzle tip has been reported in 7-94% of bottles within two weeks of use.^12,25^ Microbial cultures yield both commensal and pathogenic microbes such as *Escherichia coli*, *Staphylococcus aureus*, *Pseudomonas aeruginosa*, viruses, and fungi.^26–28^ The spectrum of identified organisms reflects the normal flora of the conjunctiva and periocular skin, suggesting contamination occurs through accidental contact with patients’ ocular surfaces or health care personnel’s hands. ^27,29–31^

In the absence of prospective clinical studies evaluating rates of infection from contaminated bottles, many hospital-based practices and surgical centers adopt self-imposed use cessation dates (SUCDs) of 1-28 days to reduce contamination risk.^10^ Despite their intention, SUCD timepoints fail to eliminate colonization risk, which can occur at any point during the use period. In a large academic practice, SUCD adherence can result in the premature disposal of 72% of total medication volume, adding $81,000 in costs per year.^32^ Recent drug shortages of essential diagnostic medications like phenylephrine and tropicamide reinforce the pressing need for practical solutions to eliminate eyedropper bottle contamination and pharmaceutical waste.^33^

Based on the current microbiological validation, the Saniteyes^TM^ system is a promising point-of-care solution for achieving fast and chemical-free HLD of reusable ophthalmic equipment. To our knowledge, this is the first technology capable of addressing both eyedropper bottle contamination and the environmental burden of single-use plastics within a single platform. The application of UVGI can realistically standardize HLD implementation across institutional and private practice settings by eliminating the operator variability and protocol inconsistencies that have historically limited compliance. This technology offers eye care providers the opportunity to enhance patient safety while reducing operational costs, staff burden, drug waste, and environmental impact. However, comprehensive clinical validation will be essential to confirm the system’s effectiveness and practical implementation across diverse healthcare environments.

## Data Availability

All data produced in the present work are contained in the manuscript.

