## Supplementary material for "Efficacy of an Automated Point-of-Care UVC Disinfection System for Reusable Ophthalmic Devices": Figures and Tables Legends

**Tables and Figures footnotes and legends**

**Table 2. Cumulative UV-C exposure by position with reference scale**

**Table 2.** UV-C colorimetric indictors were placed in three bays (rows) at four positions (columns labeled 3,6,9 and 12 o’clock) and on the nozzle tip to assess cumulative exposure. Color change reflects total UV-C dose delivered, with consistent and uniform saturation observed across all disinfection bays and positions without cold spots.

**Figure 1. Saniteyes^TM^ High-Level Disinfection System**

**Figure 1.** The ophthalmic disinfection device tested in this study (Saniteyes^TM^) utilizes three bays that can disinfect multiple ophthalmic devices in a single cycle. The tested device uses wavelengths of germicidal irradiation with primary reliance on UVC light between 254-280nm.

**Figure 2. Device Loading and UV-C Dose Validation**

**Figure 2.** Saniteyes^TM^ system with 15mL eyedroppers positioned in each disinfection bay. Colorimetric dosimeter strips were placed at 3, 6, 9, and 12 o’clock positions as well as the nozzle tip to measure cumulative exposure of high-risk contact surfaces. Pre-exposure (A) and post-exposure (B) photographs demonstrate quantitative confirmation of threshold doses of UV-C light ≥ 80 mJ/cm^2^.

**Figure 3. UVC Dosimeter Reference Card**

**Figure 3.** Intellego Technologies UVC dosimeter reference card with quantitative scale up to 100 mJ/cm^2^.
